# Cognitive Aids for Operating Room Crises – A Thematic Analysis of Implementer Experiences

**DOI:** 10.1101/2020.11.18.20234054

**Authors:** Natalie Henrich, Emily Benotti, William Berry, Alexander Hannenberg, David Hepner, Ami Karlage, Sara Goldhaber-Fiebert

## Abstract

**Background:** Strategies for the implementation of evidence-based interventions have proliferated, but it remains unclear how these strategies are operationalized for different types of interventions and contexts. Here, we examine seven implementation strategies--initially developed for implementing the World Health Organization (WHO) Surgical Safety Checklist (SSC)--for implementing cognitive aids for operating room (OR) crises.

**Methods:** We conducted semi-structured interviews with implementers of these aids exploring the use of each of the strategies previously studied with WHO SSC implementation as well as barriers and facilitators to implementation. We conducted a thematic analysis to identify and describe the use of the strategies. We mapped barriers and factors helping facilitate (facilitators) onto corresponding constructs from the Consolidated Framework for Implementation Research.

**Results:** We conducted interviews with 37 implementers across the United States. Most interviewees identified similar strategies in their implementation process, and none offered additional strategies that fell outside existing categories. There was substantial variation among implementers in how the strategies were deployed. Many of the barriers and facilitators were common across implementations.

**Conclusion:** Interviewees used a core set of strategies to implement cognitive aids for OR crises, but there was substantial variation in how implementers used these strategies, suggesting the flexibility of the strategies and the value of deliberately adapting strategies to local context. The transferability of implementation strategies from the WHO SSC to other OR-based cognitive aids demonstrates the utility of informing novel implementations with prior successful ones that share similar attributes with respect to intervention and/or context.

## Introduction

Moving evidence-based interventions into routine clinical practice is challenging. While implementation science and associated fields have produced many frameworks, theories, and models^1^ and over seventy implementation strategies,^2^ we still do not have a good understanding of how to best choose or adapt implementation efforts to the intervention at hand.^3^

Cognitive aids for operating room (OR) crises--such as the Emergency Manual: Cognitive Aids for Perioperative Critical Events, developed by Stanford Anesthesia Cognitive Aid Group (Stanford, CA) and the OR Crisis Checklists, developed by Ariadne Labs (Boston, MA)--can decrease errors and omissions during management of an OR crisis.^4^ However, given the resistance of surgical culture to cognitive aids, the long-established routines of the OR, and the infrequent (but inevitable) nature of OR crises, simply distributing cognitive aids for OR crises is insufficient to ensure their consistent and appropriate use.^5-7^ In 2015, an Anesthesia Patient Safety Foundation expert conference included a survey in which 96% of attendees agreed that “there are many steps between an individual downloading a useful emergency manual and an institution effectively implementing it clinically,” yet there was no consensus on which steps were key and how best to perform them at various types of facilities.^8^

To better understand the implementation of cognitive aids for OR crises, the research team conducted a survey of OR Crisis Checklist/Perioperative Emergency Manual implementers, asking them to select which of seven implementation strategies they used in their implementations: forming a multidisciplinary team, customizing the cognitive aid for the local facility, presenting the aid, pilot testing the aid, providing initial training, delivering ongoing training, and monitoring the use of the aid. We based these strategies on a decade of experience implementing the World Health Organization (WHO) Surgical Safety Checklist globally.^9-11^ From this survey, we found that performing more implementation strategies correlated to more reported use of the aid, but the survey did not explore how the strategies were employed.^12^

In this subsequent qualitative study, we analyzed semi-structured interviews conducted with a sample of the survey respondents selected from diverse settings, to explore how they pursued these seven strategies in their implementations. The study was undertaken to inform the enhancement of implementation advice for cognitive aids for OR crises and other similar interventions by revealing points of consistency and variation.

## Methods

### Study Design

We conducted semi-structured interviews with perioperative clinicians in the United States (US) who had implemented the OR Crisis Checklists or the Perioperative Emergency Manual in their health facility (referred to hereafter as “implementers”).

### Participants

We selected implementers from those who had completed the OR Crisis Checklist/Perioperative Emergency Manual Implementation Survey,^12^ had indicated on the survey that we could contact them for an interview, provided contact information that enabled us to recruit them via email, and came from US facilities. To increase diversity in our sample, we developed a sampling grid based on facility size (number of operating rooms (ambulatory surgery centers (ASCs) and small hospitals: 1-4 ORs, medium hospitals: 5-15 ORs, large hospitals: 16+ ORs), geography (by state), level of implementation success (based on their response to the survey item “At my facility, the tool is used regularly during applicable clinical events.”), and academic/non-academic facility. Results were analysed by facility size and level of implementation success. Geography and academic status were used to increase the diversity of the sample but characteristics were not used to stratify the results. We also conducted ten key informant interviews with individuals identified as having expertise or knowledge in a particular area related to successful implementation of cognitive aids for OR crises. Overall, we aimed for a sample size of 40 participants based on the rule of thumb that saturation (i.e. no longer obtaining new, meaningful insights)^13^ is typically achieved with a sample of approximately 30 people.^14^ Interviewees did not receive any remuneration for their participation.

### Interview Guide

We designed the interview guide to gain insight into how implementers performed the seven implementation strategies included in the OR Crisis Checklist/Perioperative Emergency Manual Implementation Survey. To determine the full range of strategies used, we also asked about any other activities conducted as part of the implementation and probed explicitly on strategies for increasing buy-in. Based on our own implementation experiences, consultation with the Emergency Manuals Implementation Collaborative (EMIC) steering committee (meeting presentations and personal communications), and implementation frameworks,^2,15^ we recognized the importance of buy-in for successful implementation. While the survey included strategies that contribute to increasing buy-in from leadership and clinicians, often as a secondary benefit, we aimed to identify additional strategies that were used specifically for this purpose. We also asked questions about barriers and facilitators to each implementation strategy. The interview guide is provided in Appendix 1.

To assess clarity and appropriateness of interview questions, we piloted the interview guide with three survey respondents and made minor changes based on the implementers’ feedback.

### Data Collection

Four team members conducted interviews, three of whom were experienced interviewers (see Appendix 2 for a list of which team members participated in each aspect of the study). An anesthesiologist-implementer from the team joined the first two interviews to ensure that the interviewers were probing the appropriate content. The senior qualitative scientist on the team monitored a sample of all the interviews to provide feedback to other interviewers and to ensure quality control as well as provided training to the novice interviewer. The novice interviewer, a prominent anesthesiologist and former president of the American Society of Anesthesiologists, may have been known to interviewees (many of whom were anesthesiologists), and he received guidance on techniques to minimize his potential influence over interviewee answers. The other interviewers had no relationship to the interviewees.

We interviewed implementers once, and interviews were approximately 45 minutes; all interviews took place between June and August 2016. Interviews were conducted by phone, audio recorded, and transcribed by an external transcription company. Members of the study team reviewed transcript quality, and as a final quality check, coders referred to the original audio recording for any necessary clarification. We removed interviewees’ names and institutions from all transcripts and stored the transcripts on a secure computer drive.

### Data Analysis

We performed a thematic analysis of the interviews in four steps. First, we coded the interviews. Study team members, including the interviewers, co-investigators, and principal investigator, created a deductive list of themes based on evidence in the literature and the interview guide. Three study team members coded the interviews in NVivo (Version 11.4.2 for Mac, Australia). The qualitative research specialist reviewed every interview’s coding to ensure quality. Throughout coding, the coders discussed any inconsistencies in themes until they reached consensus, including clarifying theme definitions as necessary.

Second, two of the coders analyzed the coded interviews thematically, focusing on how implementation strategies were used and barriers and facilitators associated with each strategy. Key findings were summarized for each theme.

Third, the analysts discussed findings with the study team to ascertain the relevance and potential implications of the findings.

Fourth, for any barrier or facilitator mentioned by at least 10% of interviewees, two team members mapped the factor onto the domains and subdomains of the Consolidated Framework for Implementation Research (CFIR).^15^ The entire study team reviewed the mapping and discussed disagreements until consensus was reached. The team selected a 10% cut off for inclusion of barriers and facilitators in order to balance the need to include factors that are likely to be relevant for other implementers while still capturing the diversity of experiences.

### Ethics

The Partners HealthCare and Stanford Institutional Review Boards reviewed the study and determined that this project meets the criteria for exemption 45 CFR 46. We obtained verbal consent before the start of every interview, with this consent method approved by both IRBs for implementer phone interviews.

## Results

### Facility Characteristics

A total of 37 implementers from 37 facilities in 19 states (Appendix 3) participated in the study. Subgroup sizes varied due to interview response rates (Table 1). No discernible differences were detected in how low-success versus high-success implementers operationalized strategies nor in the barriers and facilitators they experienced. Differences between smaller and larger facilities are indicated.

**Table 1.** Facility characteristics, by level of implementation success

|  | Non-academic facility | Academic facility | Total |
| --- | --- | --- | --- |
| **High-Success Implementers** |  |  |  |
| Small/medium hospital or ASC<sup>a</sup> (1-15 ORs<sup>b</sup>) | 8 | 2 | 10 |
| Large hospital (16+ ORs) | 1 | 17 | 18 |
| **Total** | **9** | **19** | **28** |
| **Low-Success Implementers** |  |  |  |
| Small/medium hospital or ASC (1-15 ORs) | 4 | 1 | 5 |
| Large hospital (16+ ORs) | 1 | 3 | 4 |
| **Total** | **5** | **4** | **9** |
| **Overall Total** | **14** | **23** | **37** |

<sup>a</sup> ambulatory surgical center
<sup>b</sup> operating rooms

### Implementation Strategies and Strategy Variants

Implementers reported using five strategies most frequently: forming a multidisciplinary implementation team, customizing the cognitive aid, presenting the aid, initial training, and ongoing training with the aid. Approximately half of implementers reported using either formal or informal approaches to monitor use, and only a very few formally piloted the aid prior to rolling it out across the department. Implementers did not offer implementation strategies aside from the proposed seven and the overarching category of increasing buy-in.

The ways in which implementers used these strategies (strategy variants) differed substantially (see Table 2 for all the strategy variants and illustrative quotes). For example, strategy variants for presenting the aid ranged from low effort (sending emails about the aid) to high effort (one-on-one conversations, presentations at multiple meetings). Similarly, strategy variants for customization ranged from low-tech (laminated paper copies of the aid) to high-tech (electronic versions of the aid), and strategy variants for forming multidisciplinary teams differed by number and variety of professions involved.

**Table 2.** Implementation Strategy Variants

| **Implementation Strategy**<br># of sites that discussed strategy (by facility size) | **At Any Size Facility**<br>[Variant used in both large and small/medium facilities OR variant used in only 1 facility regardless of size] | **At Large Facility<sup>a</sup>**<br>[Variant used in at least 2 large facilities and <u>no</u> small/medium facilities] | **Illustrative Quotes** |
| --- | --- | --- | --- |
| **Form a multidisciplinary team**<br>Total n=12/28<br>Small/medium n=3/10<br>Large n=9/18 | Professions included on teams:<br>• Anesthesiologists<br>• Nursing<br>• patient safety/quality improvement leaders<br>• OB/GYN*<br>• Information technology*<br>• Design and innovation*<br>• CRNAs*<br>• Residents*<br>• Surgical education colleague*<br>Function of team:<br>• Help with customization<br>• Review the aid* | Additional professions included on teams:<br>• Pharmacists<br>• Surgeons<br>Additional functions of team:<br>• Used the team to help with implementation plan (in addition to customizing) | “You really want interdisciplinary conversation, implementation, and practice from the get-go. The day that one person manages the emergency in an OR is the day that emergency won’t go very well. And you need everybody. So, let’s be colleagues and let’s be a team. Let’s talk about how we’re going to implement it, revise it and make it work so everybody at the end of the day knows it is better. And in doing so, you will find out that your nurses are your best friends.” (Large)<br>“It went through various committees. We have a patient safety committee that is made up of interdisciplinary team members. We have an OR executive committee so we had buy-in at each level so it wasn’t a project that was done solely spearheaded by one department which, in our experience, always leads to a project failure.”(Large) |
| **Customize**<br>Total n=18/28<br>Small/medium n=6/10<br>Large n=12/18 | Changes to content:<br>• Added or subtracted protocols to fit facility<br>• Added local phone numbers<br>• Ensured pharmacy formulating was consistent* | Additional changes to content:<br>• Made own manuals informed by existing manuals<br>Additional customizations: | “I think [customizing] helped with the success of the checklist because the people that worked on either the customizations or owned that little part felt that they were the content expert. We felt local buy-in so the checklists are branded like our institutions. They are customized with phone numbers. They are customized with workflow and |
|  | Other customizations:<br>• Laminated pages<br>• Added page to explain institution's emergency call button system*<br>• Added address*<br>• Changed language from "anesthesiologist" to "crisis manager"*<br>• Used local spellings*<br>• Added tabs to each protocol*<br>• Eliminated some pages that weren't relevant to their location*<br>• Made some directions more concise*<br>Miscellaneous:<br>• Printed black and white copies to lower costs*<br>• Used a bright red binder*<br>• Laminated pages*<br>• Placed in clear plastic envelopes and velcroed to wall*<br>• Added back page that showed how to document critical events in the OR*<br>• Added flow map for emergencies* | • Created electronic format, available on local computers<br>Miscellaneous:<br>• Had multiple interdisciplinary people help with customization to increase buy-in | then we've even further customized not just our main campus but our satellite site that is six blocks away that has a little bit of a different workflow or different phone numbers or different patterns of doing things so it makes it a workable checklist that really, in a local environment, is helpful." (Large)<br>"Don't just cookie cutter, you know, take something, print and plunk and expect it to be used." (Large)<br>I took about four or five different ones and added and subtracted and pulled and removed...we took a whole bunch of things that [made sense] for our surgery center. We took things that didn't make sense and omitted them...we have a hybrid. We took Harvard's and Stanford's and then put them together. (Large) |
| **Present aid**<br>Total n=22/28<br>Small/medium n=7/10<br>Large n=15/18 | Where/how presented:<br>• Meetings (staff/department)<br>• Emails<br>• 1:1 conversations<br>• PowerPoint presentations*<br>• During orientations*<br>• Time before cases*<br>• Morning huddle*<br>• “Electronic format”*<br>• Section line meetings*<br>• Quality department*<br>• Teleconference*<br>• Newsletters*<br>• Word of mouth*<br>Who it was presented to:<br>• Anesthesiologists<br>• Nurses<br>• Surgeons<br>• Simulation educators<br>• OR administrators<br>• Key champions*<br>• Front desk staff*<br>• Surgical technicians*<br>• Circulators*<br>• OR team*<br>• PACU staff*<br>• Quality dept*<br>• Med exec*<br>Content of presentations:<br>• Rational/purpose of aids<br>• Promote awareness/familiarity | Additional places presented:<br>• Grand rounds<br>Additional people to whom it was presented:<br>• Clinicians in training (i.e., residents/students) | “[Presenting the checklist] can be done by in service, one-to-one in the operating room, discussions, things of that nature...definitely formal and informal.” (Large)<br>“We had a staff meeting...it was our administrative office staff to our nurses to - it was everybody. Doctors, nurses, everybody across the board.” (Small/medium) |
|  | • How to use the aid*<br>• What the aid covers*<br>• Provide encouragement* |  |  |
| **Pilot test**<br>Total n=3/28<br>Small/medium n=1/10<br>Large n=2/18 | Piloted in simulation center*<br>Piloted in tabletop simulation*<br>Used in drills (in-situ simulation), then modified*<br>Piloted with residents* |  | “I think a valuable piece of that step is some more testing amongst the anesthesiologists themselves, and then perhaps it's worthwhile to bring in a couple of other team members, and pick some of the checklists and actually mock-test them...So, a tabletop simulation, or you know, not sophisticated simulation in a medical simulator. Some way of testing a few of them anyway to make sure that they actually are what you want before you ask other people to use them.” (Large) |
| **Training**<br>Total n= 26/28<br>Small/medium n= 8/10<br>Large n=18/18 | Methods:<br>• Low tech training (e.g. drills)<br>• Ongoing training<br>• 1:1 peer training<br>• Debrief after training*<br>• Incorporate into orientation*<br>Clinical Participants:<br>• Nurses<br>• Anesthesiologists<br>• Educators*<br>• Perioperative team*<br>• Code blue team*<br>Non-clinical Participants:<br>• Administrative staff* | Additional methods:<br>• Used high and low tech simulations (small/medium sites only used low tech simulations)<br>Additional clinical participants:<br>• Included surgeons, surgical technicians, residents (full interprofessional OR teams) | “I’ve evolved as a simulation person, I started out, you know, everything had to be high fidelity and it’s as close to real life as possible and then now all the time I feel like you can actually get a lot more done if you just kind of keep it simple.” (Large)<br>“And then in the simulations themselves, if they’re not using the manual, we’ll throw in some prompts. I asked them, ‘Do you want to use the manual,’ or if they’re struggling with coming up with particular medications and I say, ‘Well, I think that medication is in the manual.’... It just goes to show that it is valuable and that it sort of jogs their memory to use it.” (Large)<br>“It was typically like a 20 or 30 minutes thing and what I would typically do was get one of the members in |
|  | Other:<br>• During simulations, participants also worked on teamwork and communication skills<br>• Empower nurses to speak up through training*<br>• Check-in meetings every 3-4 months to understand how training is going and address any issues* |  | the staff to pretend to be a patient. And we would make it relevant to what we did. So we are an orthopedic surgery center. So you'd say oh you know, we are doing a knee scope on this healthy athlete... And then we would start from there. I'd have somebody get the checklist and we'd bring in the code call, we'd have people just kind of go through it and then it feels like a little board exam. You know you'd have one problem turn into another problem, so you'd go from like one protocol to the next...so it's just the little scenarios that I basically made up." (Small/Medium) |
| **On-going training**<br>Total n=14/28<br>Small/medium n=5/10<br>Large n=9/18 | Methods:<br>• Simulation (including drills/in-situ simulation)<br>• Incorporate into Health Streams/courses<br>• Incorporate into M&M meetings*<br>Frequency:<br>• 2/year<br>• 1/year<br>• Every 2 years<br>• Ongoing during residency*<br>• Quarterly*<br>Who gets trained:<br>• New providers<br>• Residents<br>• All staff |  | “Periodic drilling, I think is the obvious answer...I've been around the simulation community for a long, long time, and people ask when they come into the simulator, "How often do I need to be refreshed?"...I think that you need to keep practicing with it over time if we're going to turn it into a norm. Right? I think that that's what [happened] in the places that really do the surgery safety checklist well. If I went in and took their checklist away from them, everyone in the room would feel uncomfortable doing the next case. How long does it take to get there? How many uses of it does it take? I don't think anybody's studied it.” (Large)<br>“It may sound like we're beating a horse to death but what we would do is that we would have one of our M and M conferences and if the M and M involved one of the crisis checklist items then we would take that opportunity to reinforce to everybody what you would do if that happened during the case. So again, it brings it up to them, so<br>if someone presents a case of venous air embolism, then we always have to take that opportunity to say, “Okay, and remember if you have a venous air embolism this is how you access the crisis checklist and these are the tenets of management based upon the crisis checklist.” (Large) |
| **Monitor use**<br>Total n=13/28<br>Small/medium n= 5/10<br>Large n=8/18 | Informal monitoring mechanisms:<br>• Champion asks if people have used it; records stories of use<br>• Staff shares anecdotes of use with champion<br>• Champion asks about use during casual conversations<br>Formal monitoring mechanisms:<br>• Survey*<br>• Weekly check-in with anesthesia scheduler who runs desk*<br>• Paper log of emergencies and indicate in log if aid was used* | Formal monitoring mechanism:<br>• EMR data<br>• Review use of aid in post-crisis debrief | “I would query the staff and say, ‘has anybody used it?’, ‘did anybody even pull it out and take a look at it?’ (Small/medium) |

Notes:
<sup>a</sup> There were no strategies mentioned exclusively by small/medium facilities [i.e., used in at least 2 small/medium facilities and no large facilities].
\* indicates the strategy variant was used in only 1 facility (n=1).

Within this variation, large facilities generally pursued strategies in ways that reflected both their larger bureaucracy and greater resources. At large facilities only (defined as reported by 2 large facilities and <u>no</u> small/medium facilities), strategies such as customization, presenting the aid, and training included more participants from a wider range of professions and residents. Similarly, only large facilities implemented both electronic and printed versions of the aid and/or trained clinical staff using high-fidelity simulations as well as presentations. The most notable distinction between large and small/medium facilities was in monitoring. Six large facilities reported using formal monitoring mechanisms with 4 strategy variants; in contrast, only 1 small/medium facility reported using a formal monitoring mechanism. No strategy variants were reported exclusively by small/medium implementations (defined as reported by 2 small/medium facilities and <u>no</u> large facilities).

When asked about increasing buy-in, implementers identified three main strategy variants: 1) having one-on-one conversations with leadership and resistors, 2) presenting the aid at meetings, and 3) using the aid in simulation/training drills. These variants served multiple purposes: in addition to motivating the use and spread of these aids, one-on-one conversations and presenting the aid at meetings are both methods for making people aware of the aids and their purpose. Simulations/drills are ways to train OR teams on the use of the aid while also showing their purpose in a powerful experiential way. One implementer described the reaction of residents managing a crisis with and without the manual as “eye-opening…people are like ‘Wow! That was so much easier with this manual in place. It really helped me through the treatment process.’” She emphasized that the best way to achieve this eye-opening effect was through direct experience in simulation training.

At times, implementers involved leaders and other non-clinical critical staff members in implementation strategies that were developed for clinicians as a way of increasing buy-in across stakeholder groups. An implementer who customized the aid for use in an office setting reported she “felt like it was important for [the office manager] to be [at the training] because she might be the one calling the ambulance…one of my [locally added] protocols is the transfer of a patient to the hospital, which numbers do you call, who’s responsible for calling 911, stuff like that…and being part of the surgery center administrative support is important, so she was involved.”

### Facilitators and Barriers

A small number of facilitators (Table 3) and barriers (Table 4) were common across many of the implementing sites. Buy-in was the most commonly cited facilitator, with nearly half of implementers expressing that general support from colleagues facilitated implementation. Some implementers specifically mentioned that support from the chair of anesthesia, nursing leadership, and/or the OR/perioperative director were useful. Implementers also indicated that clinicians’ acceptance of cognitive aids (i.e., having positive views of cognitive aids in general, not just this one) facilitated implementation. Several respondents spoke of the importance of changing organizational culture to make the use of cognitive aids in clinical work more acceptable. To begin this cultural shift, they recommended having providers participate in simulations with and without the aid to experience the difference in performance with the cognitive aid (training). Overwhelmingly, the most commonly cited barrier was insufficient time for training which then made scheduling difficult, especially for attending physicians. Conversely, dedicated time for training was frequently mentioned as a facilitator.

**Table 3.**
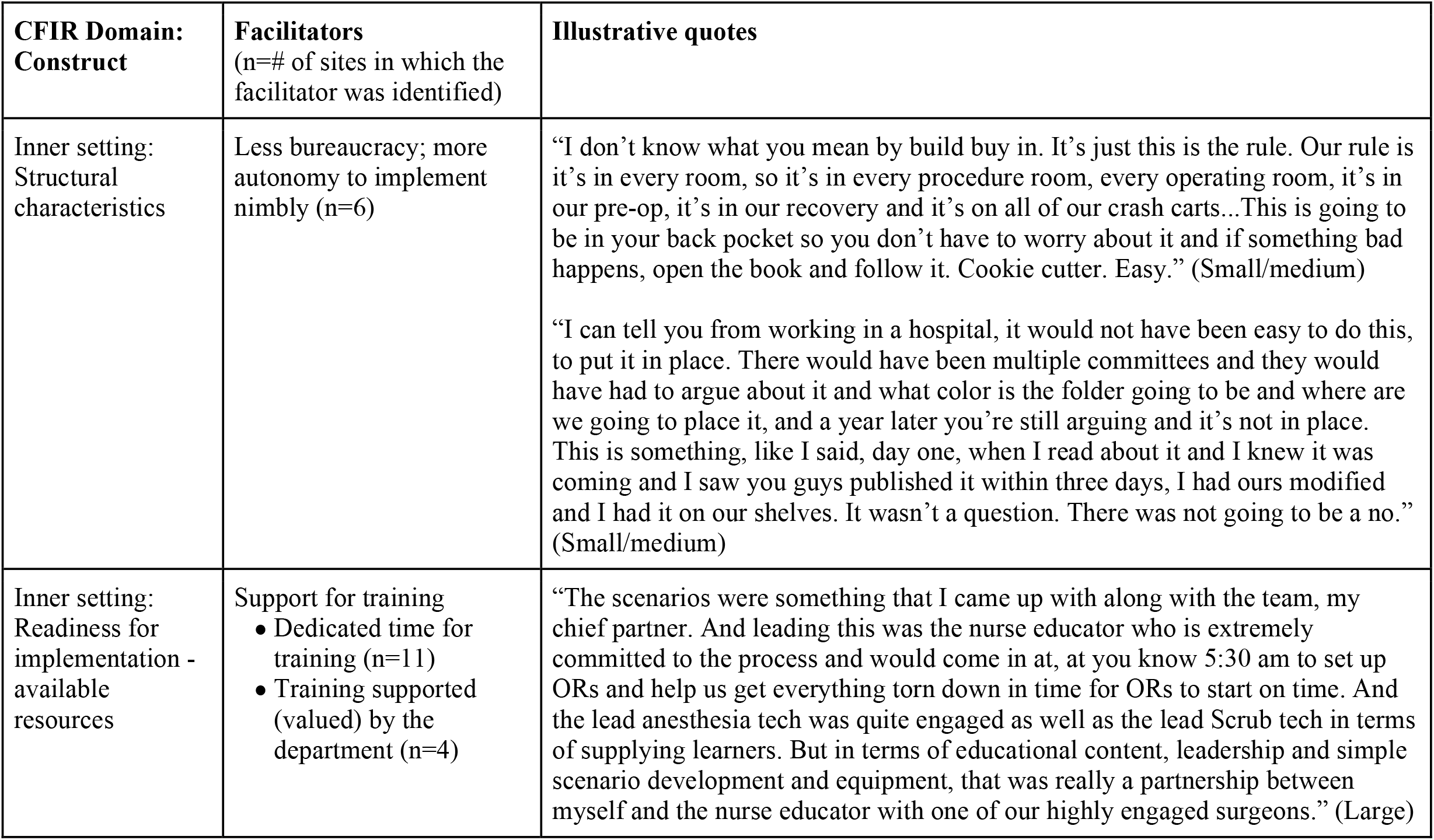
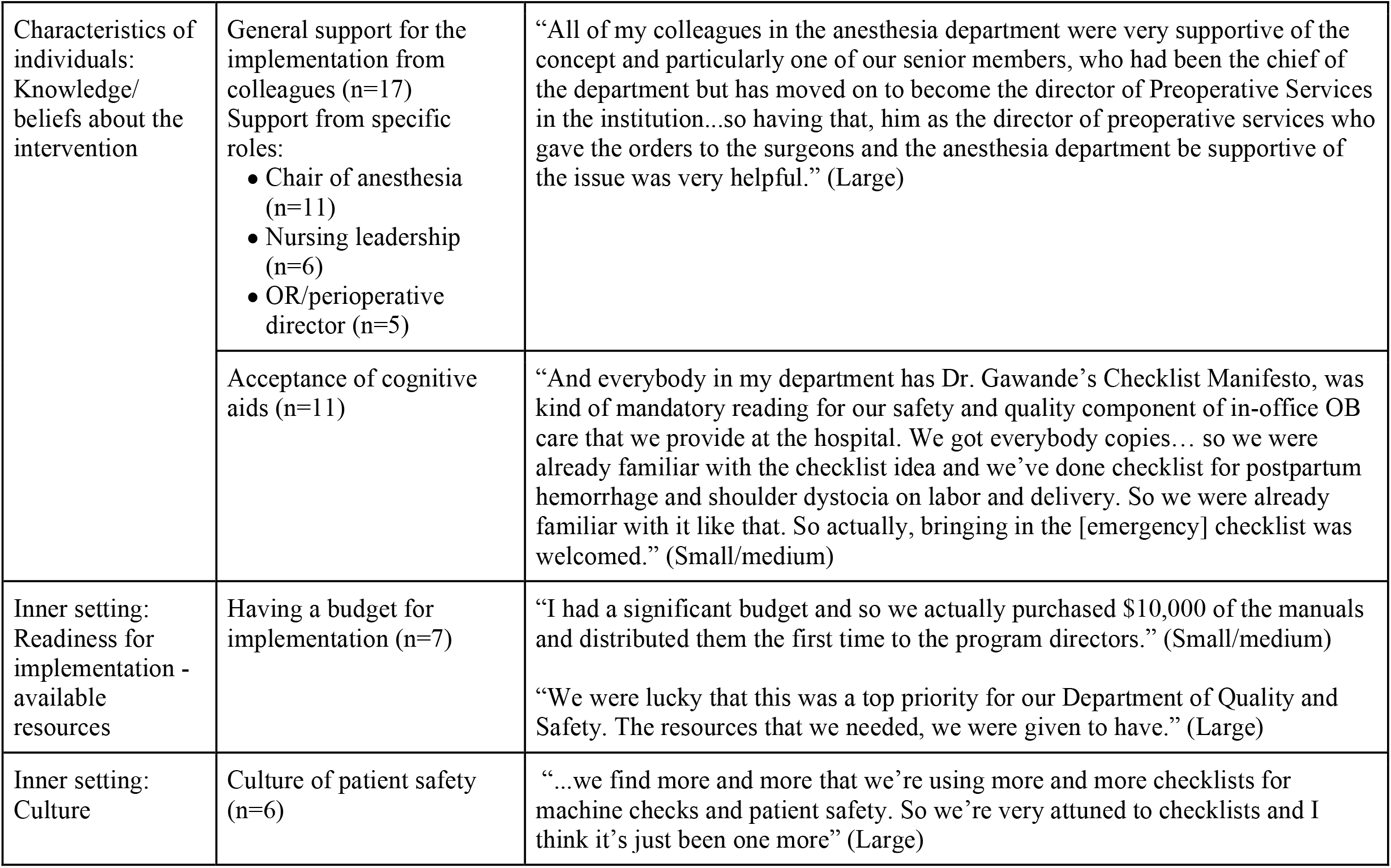
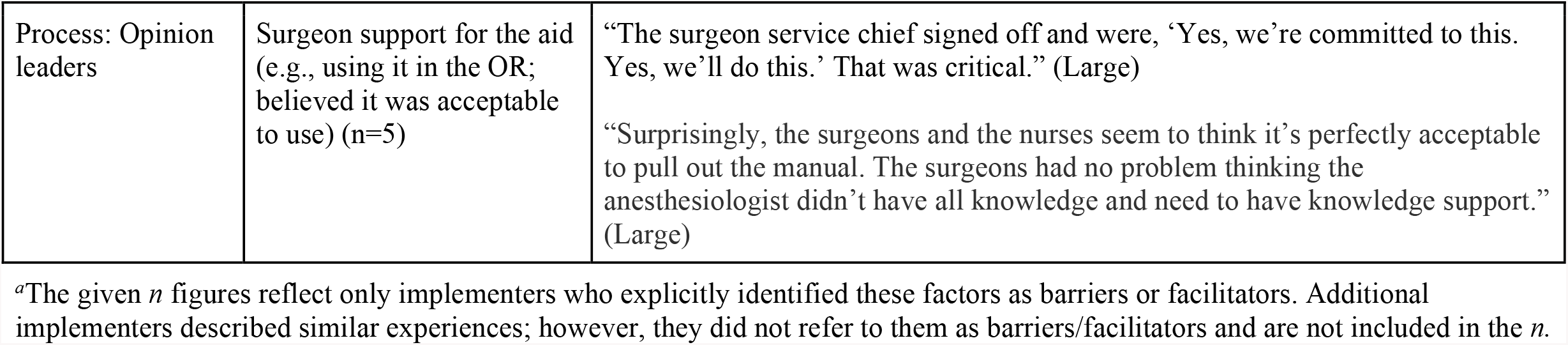
Facilitators Mentioned by at Least 10% of Implementers, by Consolidated Framework for Implementation Research (CFIR) domains^*a*^.

**Table 4.**
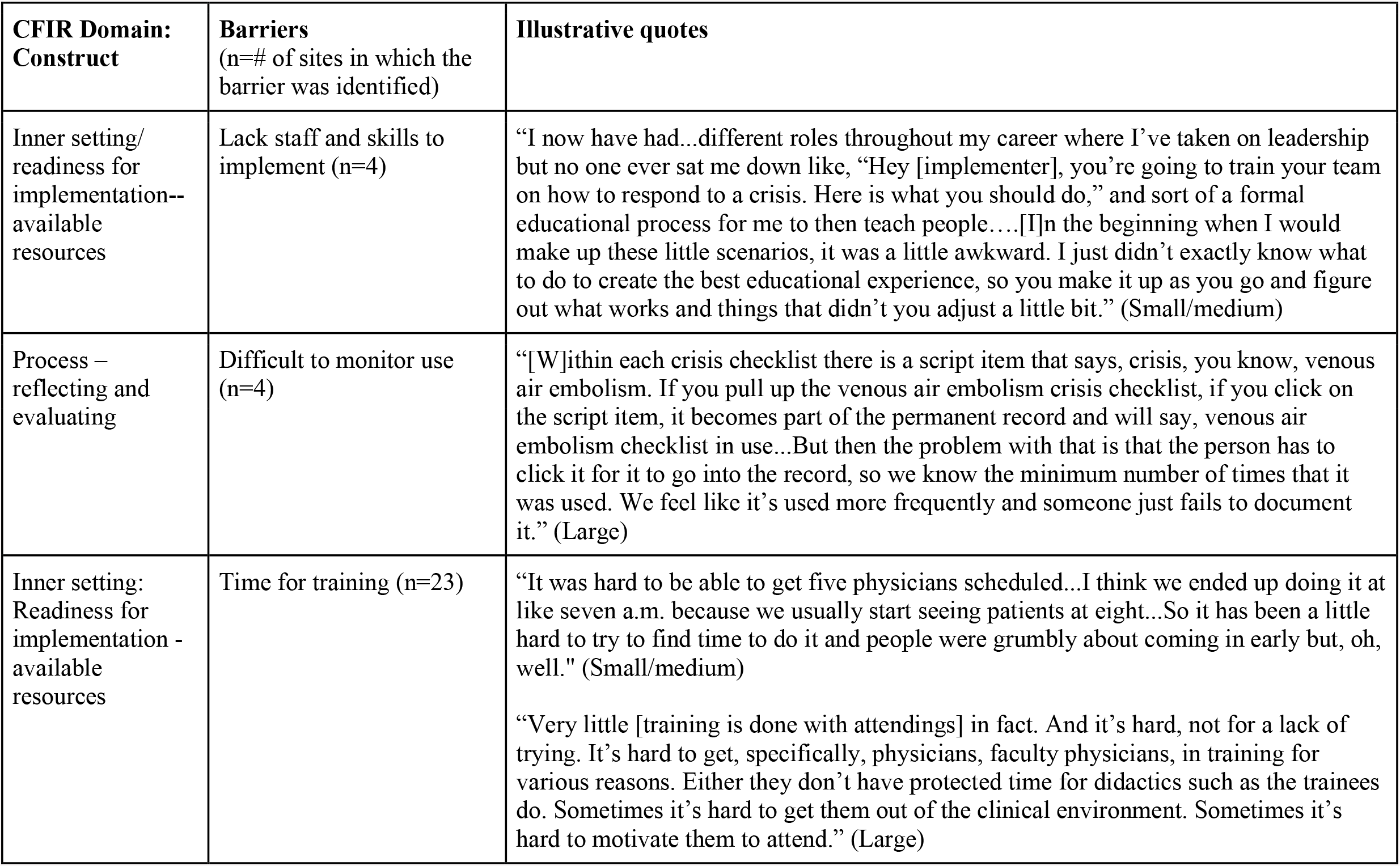
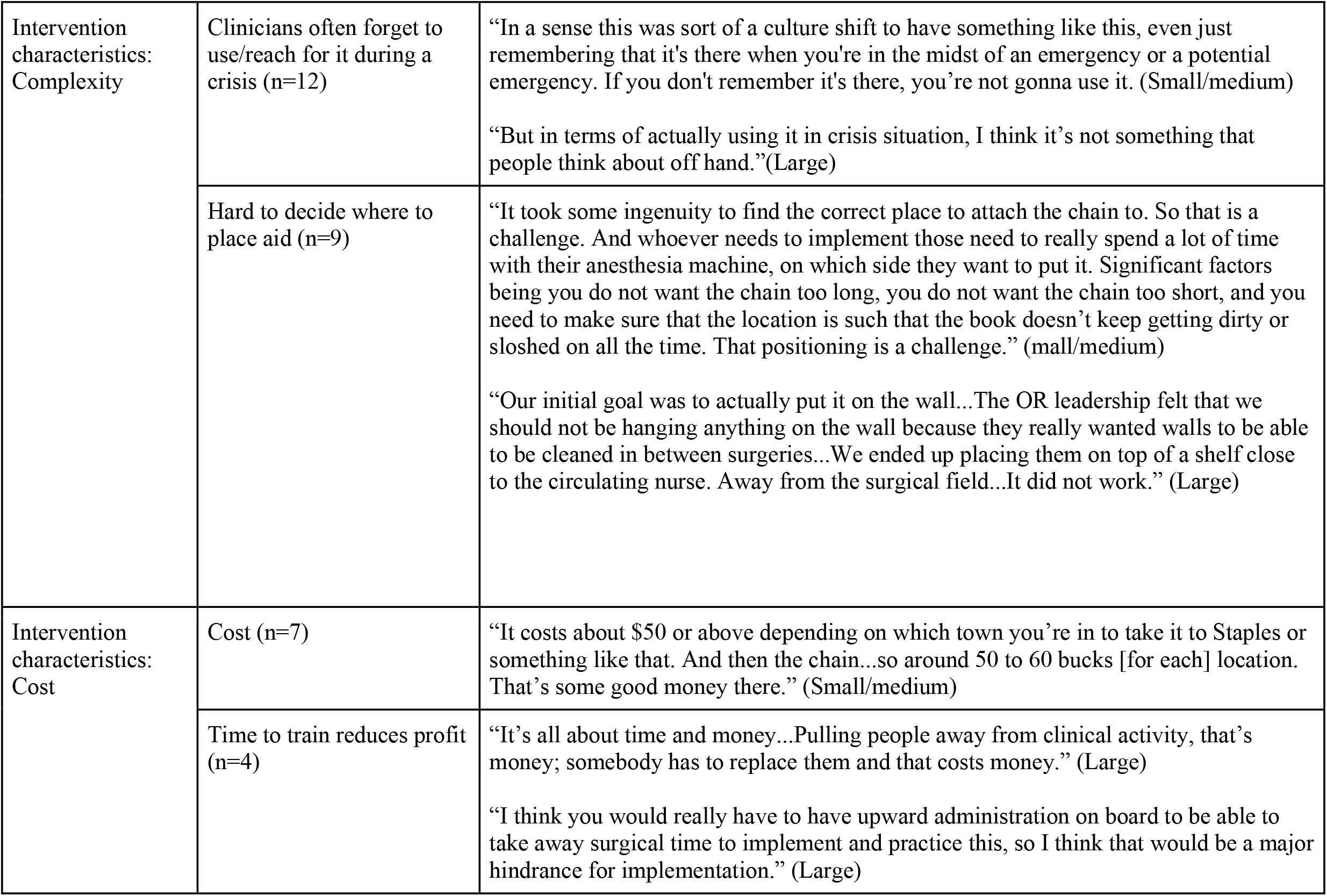
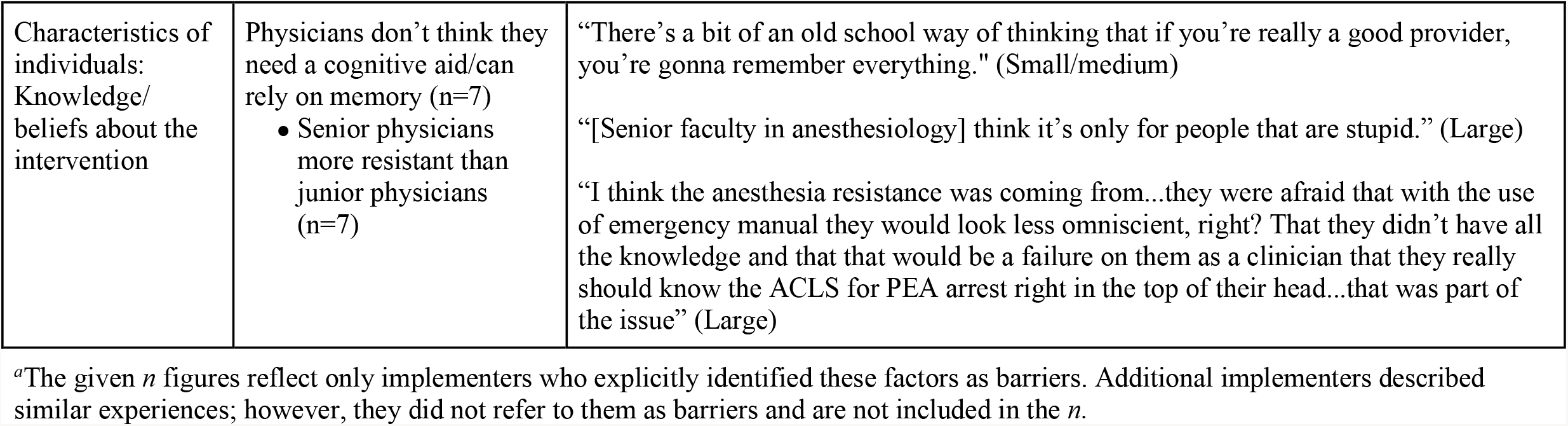
Barriers Mentioned by at Least 10% of Implementers, by Consolidated Framework for Implementation Research (CFIR) domains^*a*^.

Some barriers and facilitators clustered by facility size. Small/medium facilities provided more opportunity to implement nimbly with less bureaucracy, increasing the efficiency of the implementation and enabling a single implementer to lead with minimal coordination or permissions required. For instance, while an implementer in a small/medium facility might hold a one-on-one conversation over a happenstance lunch with a single “influential” colleague to increase buy-in, an implementer in a large facility might need to schedule a lunch in advance with three “influential” colleagues in order to achieve the same effect. Simultaneously, the reduced size of small/medium facilities also created barriers: lack of staff to support implementation, implementation expertise, and other resources led to a heavy dependence on a single implementer, which in turn could impede implementation.

In contrast, large facilities provided access to more staff, resources, and infrastructure for implementation. Implementers from large facilities spoke about intensive efforts for integrating formal training with the aid, with more resources and skilled leaders available for single or interprofessional immersive trainings. Some implementers found it helpful to think creatively about scheduling trainings during less busy clinical times, such as protected educational times, early mornings, weekends, and slower days. Simultaneously, large facilities suffered from increased bureaucracy, including slower and more formal change processes, complex scheduling barriers, and the need for many one-on-one conversations and large meetings to increase buy-in. For implementers from large facilities, monitoring the use of the aid during relevant clinical events posed a notable challenge. They found it impossible to reach all clinicians through informal monitoring mechanisms, and system constraints limited formal attempts to monitor use of the aid via the EMR. Monitoring challenges included uncertainty around why the aid was accessed (e.g. educational purposes or responding to a crisis), knowing only the crises in which the aid was used but not the crises in which it was not used, and incomplete clinician documentation.

## Discussion

We conducted a qualitative analysis of semi-structured interviews with implementers of cognitive aids for OR crises in which we inquired about seven implementation strategies adopted from the Surgical Safety Checklist implementation guide and about strategies for generating buy-in. No facilities reported strategies outside these seven strategies or the overarching buy-in category, although the ways people implemented these strategies (strategy variants) differed significantly. These implementation strategies are therefore likely to be sufficient for facilities seeking to implement the cognitive aids for OR crises, however, some of these strategies, and ways of applying them, are likely more important than others, at least in some contexts. For example, only 3 facilities pilot-tested the aid (i.e., small scale use of the aid prior to full roll-out) and all of these were conducted in simulations. Pilot testing of these aids by other means is difficult because the need to use them is rare given the low occurrence of crises, unlike for the WHO Surgical Safety Checklist which is used in every operation.^16^ In contrast, buy-in appears to be an essential component of implementation that gets infused into multiple strategies and is fostered both in activities that are explicitly aimed at generating support, as well as in strategies designed to serve other implementation objectives, such as training. Future research on the relative importance of strategies and ways of applying them, in various settings, would be a valuable contribution to the field.

The diversity in strategy variants suggests that implementers can utilize these seven implementation strategies in many different ways. Moreover, while we were not able to link specific variants to successful or unsuccessful implementation of the cognitive aid for OR crises, we believe that deliberate adaptation of an implementation strategy to a specific context likely leads to stronger implementation. For instance, large facilities frequently suffered from the barrier of increased bureaucracy, but implementers were able to utilize the strategy of presenting the concept to multiple important stakeholders in order to move the implementation process forward. Similarly, in small/medium facilities with less bureaucracy but also fewer resources, implementers were able to utilize one to one conversations to generate crucial buy-in for the aid and commitment to implementation from clinicians. Thus, much like the aids themselves, implementers ought to carefully customize implementation strategies to their context.

Our results also suggest that implementation strategies developed or chosen for one intervention (the WHO Surgical Safety Checklist) can be successfully adapted to similar interventions (cognitive aids for OR crises). We remain uncertain as to what factors are most important in judging transferability a priori. Here, it may be the style of intervention (cognitive aid), the context (OR), or the people involved (surgical teams). As with the relative importance of strategies, determining transferability is another area ripe for future research.

We found that some barriers and facilitators varied in prevalence by facility size. However, implementation success may depend less on the size of the facility than on implementers’ adaptations to that size, harnessing inherent strengths and mitigating weaknesses. Size is only one of many organizational context characteristics implementers should consider as part of a deliberate adaptation of strategies for their facility. As identified in the CFIR framework, implementers may also need to consider other inner setting (local) factors, outer setting (context external to the organization) factors, the individuals in the organization, and implementation process, in addition to intervention characteristics. ^15^

Our study has several limitations. The final sample contained somewhat less variation in facility characteristics than we intended. However, it included geographical diversity with interviewees from 19 states and representation of every cell in our demographic sampling schema, suggesting that our respondents were sampled from across a wide spectrum of implementation contexts despite the deviation from the original sampling plan. We are also unable to determine which strategy variants correlate to successful or unsuccessful implementation. We reached saturation in the implementation strategies as well as for barriers and facilitators, as we were no longer hearing novel variants. However, it is possible that we did not detect other rarely used variants or other rarely occurring barriers and facilitators. This is a limitation of all qualitative research, which is not intended to be generalizable but rather to reflect a sampling of the range of variation in a population.

## Conclusion

This qualitative analysis of interviews with implementers of cognitive aids for OR crises demonstrates that the seven recommended implementation strategies, based on past experience with the WHO Surgical Safety Checklist, are relevant but possibly not all necessary for successful implementation of cognitive aids for OR crises. Facilities achieved varying implementation success using these strategies in diverse ways, some of which corresponded to facility size. As a practical extension of this research project, a cost-free toolkit was also developed from this research, containing modifiable resources for implementation of and training with these tools, available at https://www.implementingemergencychecklists.org/.

## Supporting information

COREQ checklist_Cognitive Aids for OR Crises

## Data Availability

Data are not publicly available but can be requested by contacting the corresponding author.

## Acknowledgements

This project was funded primarily under grant number 5R18HS024235-02 from the Agency for Healthcare Research and Quality (AHRQ), US Department of Health and Human Services. The opinions expressed in this document are those of the authors and do not reflect the official position of AHRQ or the US Department of Health and Human Services. The funder had no role in the analysis, interpretation, or publication of this article

## Appendix 1. Interview Guide

### OR Crisis Checklist Interview

Thank you for speaking with me today about implementing the OR Crisis Checklist/Emergency Manual at your facility. You may recall that you completed our online OR Crisis Checklist/Emergency Manual Survey. There were some very interesting findings from the survey and our goal with the interviews is to better understand the survey results by more deeply exploring what people did and did not do during implementation, and why. Ultimately, this information will be used to inform the development of an OR Crisis Checklist/Emergency Manual Implementation Tool Kit that will be provided to you at the conclusion of our project.

The interview will take approximately 45 minutes and will be audio-recorded so that we can transcribe the conversation for analysis. During the transcribing process all names of people and facilities, and any other identifiers, will be removed but I’d appreciate if you could please try to avoid using names and instead identify people by their role and facilities by their type. We will keep your identity confidential and no individual people or facilities will be identified in the presentation of the results.

Do you have any questions before we begin?

1. From the time it was decided to implement the checklist/manual at your facility and throughout the implementation process, whose support was most important to your implementation?
  a. In what ways did they show their support?
2. What was done to build buy-in for using the checklist/manual?
3. There are many things a facility can do to support implementation of the OR Crisis Checklist/Emergency Manual. When you did the survey you indicated activities that were and were not done at your facility. I’d like to go through that list again so we can discuss each of the activities in more detail, including reviewing what you did and how you did, what you didn’t do and why, and how you think the activities impacted the success of implementation.

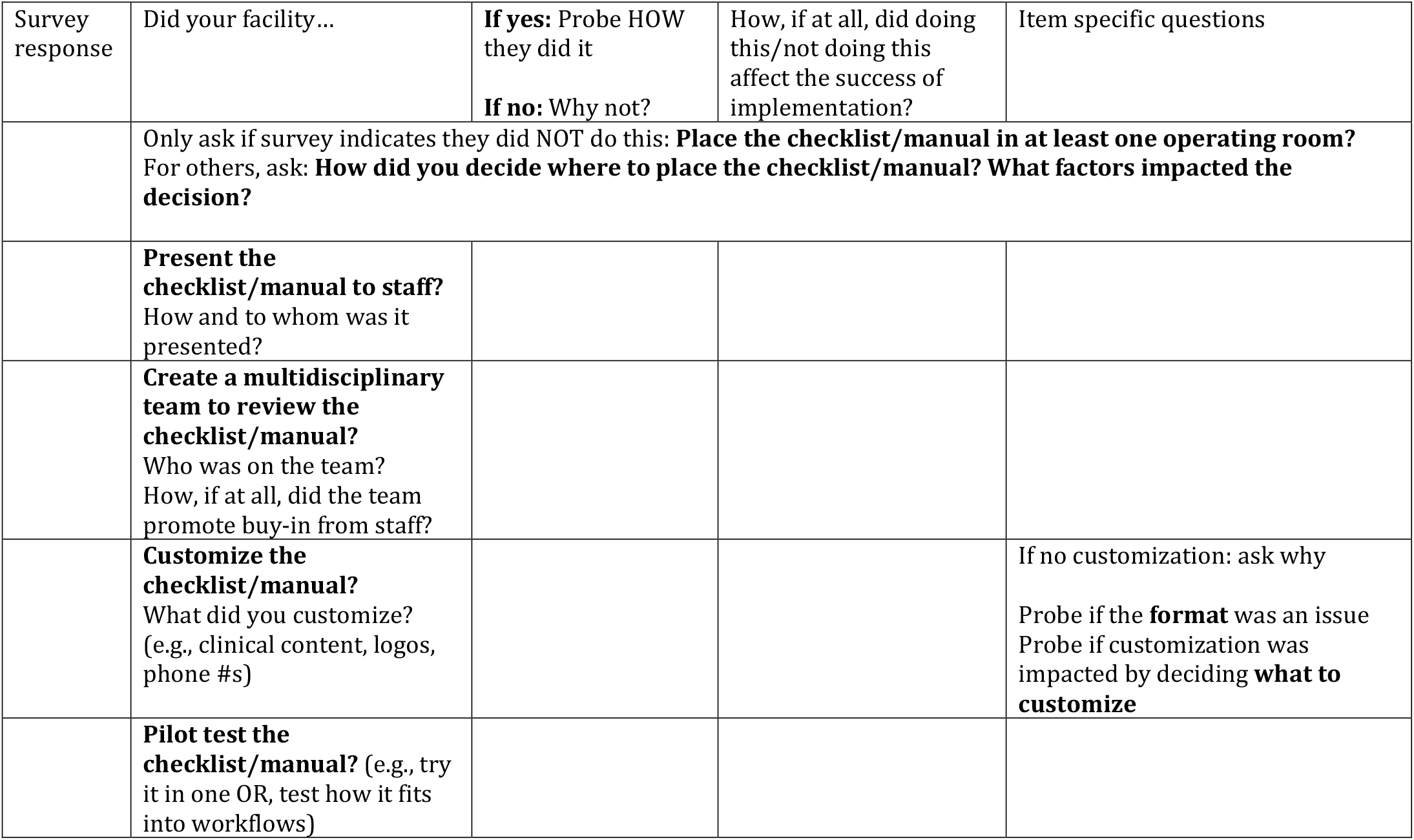
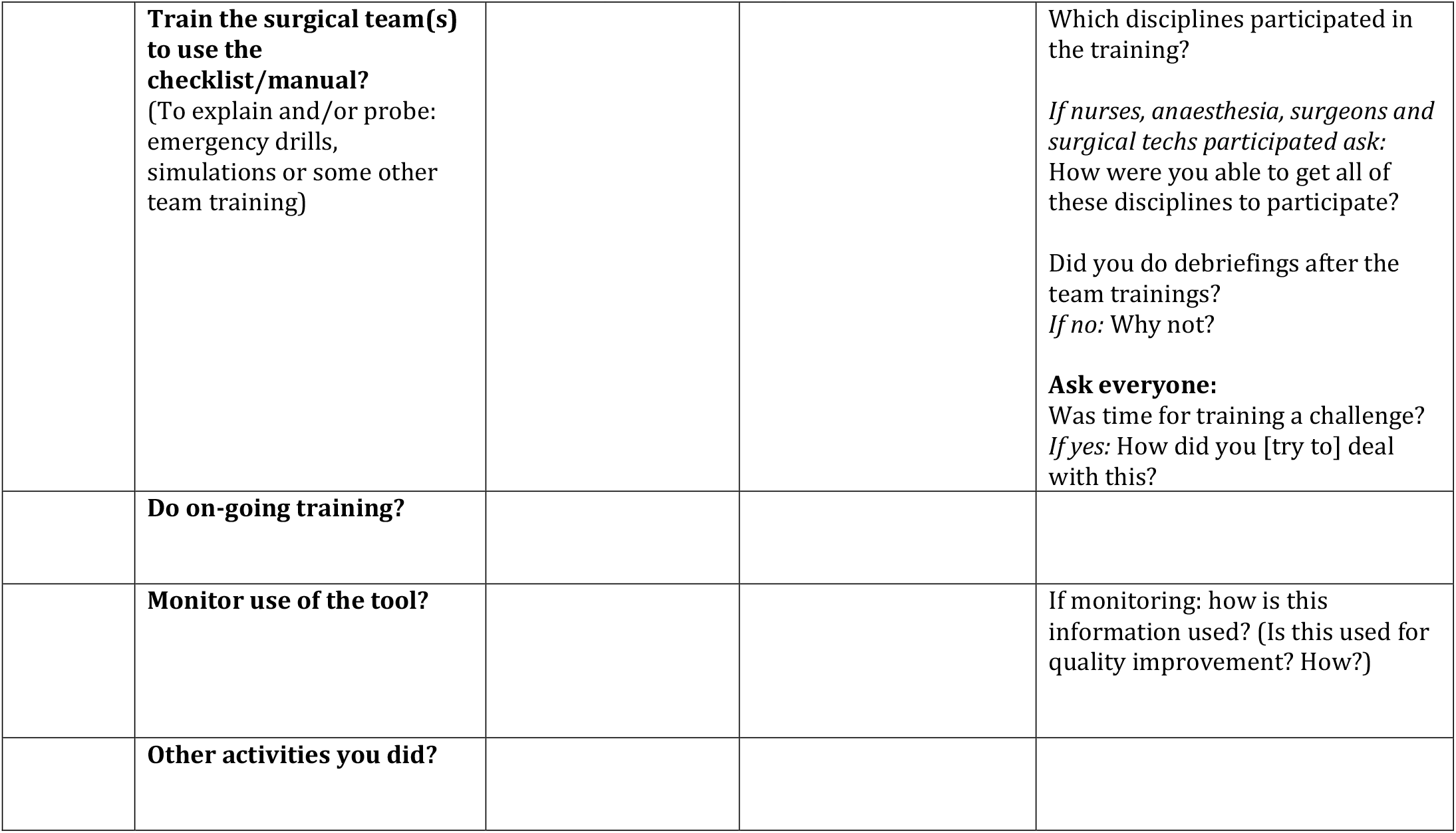

### Q4-6 IF TIME PERMITS (otherwise continue to Q7)

4. How did you decide to do these implementation activities? [refer back to what was done]
5. Are there other things you wanted to do as part of bringing the checklist into use at your facility but that you haven’t done? If yes, why haven’t you done these? (explore challenges/barriers)
6. Do you think there are other things that could be done to make use of the checklist at your facility even more successful? What are they?
7. We’re interested in understanding the challenges your facility experienced in implementing the checklist/manual, and how your facility responded to these challenges. What were the challenges you faced in getting the checklist into use?

8. Has your facility used the checklist/manual during an OR crisis?
  **If no:** Why not?
  **If yes:** Can you describe a case for which the checklist/manual was used?
  Did the checklist/manual help the clinicians involved? How?
  How successfully do you think the checklist/manual was used?
  Why do you think you had this level of success?
  **If used checklist/manual:** Did you debrief after the event?
  **If no:**
  Why not?
  Is there someone at your facility with experience as a facilitator or debriefer?
  What do you think you would have gotten out of a debrief?
  **If yes:**
  Who led or initiated the debrief? Why were they chosen?
  Who participated in it?
  What did you get out of doing the debrief?
9. If there was an applicable critical clinical event in your facility tomorrow, what percentage of the time do you think the checklist/manual would be used?
  a. Why?
  b. How would you know if it was used?
  c. What/who would trigger using the checklist/manual? (e.g., when a critical event begins, after the team tries to resolve the crisis but is unsuccessful, after the event is dealt with to make sure steps weren’t missed, in debrief)
10. Have there been any negative consequences to introducing the checklist/manual?
11. Having gone through the process of implementing the checklist/manual, what would you have done differently if you could do it all over again?
  a. What would you do the same if you could do it again?
12. What advice would you give a facility that was considering implementing the OR Crisis Checklist/Emergency Manual?
13. Is there anything else about implementing or using the checklist/manual that you’d like to mention?

| Challenges identified in survey | Challenge | How did you facility deal with [add challenge]? | How successful do you think your facility was at dealing with this challenge? | Item specific questions |
| --- | --- | --- | --- | --- |
|  | Opposition/resistance<br>[In addition to the challenges you've mentioned, was there opposition or resistance from anyone about implementing the checklist?] |  |  | From whom was there resistance?<br>Why did they resist?<br>Is it considered acceptable to use the checklist/manual? |
|  | If **budget** is a challenge:<br>What helped/hindered you getting a budget?<br>What information would help you get your administrators to give you an appropriate budget? |  |  |  |

## Appendix 2. Authors’ contributions

Natalie Henrich: This author helped with study design, data collection, analysis, and writing of the manuscript.

Emily Benotti: This author helped with data collection, analysis, and writing of the manuscript.

William Berry: This author helped with study design, analysis, and writing of the manuscript.

Alexander Hannenberg: This author helped with study design, data collection, analysis, and writing of the manuscript.

David Hepner: This author helped with study design, analysis, and writing of the manuscript.

Ami Karlage: This author helped with analysis, and writing of the manuscript.

Sara Goldhaber-Fiebert: This author helped with study design, data collection, analysis, and writing of the manuscript.

## Appendix 3. Participants’ State

1. California
2. Colorado
3. Delaware
4. Florida
5. Illinois
6. Iowa
7. Maine
8. Massachusetts
9. Michigan
10. Minnesota
11. New Jersey
12. New York
13. North Carolina
14. Oregon
15. Pennsylvania
16. Rhode Island
17. Tennessee
18. Vermont
19. Washington

