## Supplementary material for "Cognitive Aids for Operating Room Crises – A Thematic Analysis of Implementer Experiences": COREQ checklist_Cognitive Aids for OR Crises

Consolidated Criteria for Reporting Qualitative Studies (COREQ): 32-item Checklist

Manuscript Title: Cognitive Aid for Operating Room Crises - A Thematic Analysis of Implementer Experiences

| No. Item | Guide questions/description | | Location |
|---|---|---|---|
| **Domain 1: Research team and reflexivity** | | | |
| *Personal Characteristics* | | | |
| 1. Interviewer/facilitator | Which author/s conducted the interview or focus group? | Natalie Henrich, Emily Benotti, Alexander Hannenberg | Appendix |
| 2. Credentials | What were the researcher's credentials? E.g. PhD, MD | Natalie Henrich, PhD, MPH; Emily Benotti, MPH; William Berry, MD, MPA, MPH; Alexander Hannenberg, MD; David Hepner, MD, MPH; Ami Karlage, BA; Sara Goldhaber-Fiebert, MD | Authors page |
| 3. Occupation | What was their occupation at the time of the study? | The researchers' occupations include: research scientists/faculty, research specialist, physicians/faculty, writing specialist | -- |
| 4. Gender | Was the researcher male or female? | Interviewers included males and females | -- |
| 5. Experience and training | What experience or training did the researcher have? | Experts in anesthesiology, cognitive aids use, implementation science, and qualitative research | -- |
| *Relationship with participants* | | | |
| 6. Relationship established | Was a relationship established prior to study commencement? | One interviewer might have been known to participants | Methods |
| 7. Participant knowledge of the interviewer | What did the participants know about the researcher? e.g. personal goals, reasons for doing the research | Participants might have known one interviewer through anesthesiology networks | Methods |
| 8. Interviewer characteristics | What characteristics were reported about the interviewer/facilitator? e.g. Bias, assumptions, reasons and interests in the research topic | Levels of experience of interviewers; possible relationship between one interviewer and participants and that | Methods |

| | | | |
|---|---|---|---|
| | | interviewer's role outside of this project | |
| **Domain 2: study design** | | | |
| *Theoretical framework* | | | |
| 9. Methodological orientation and Theory | What methodological orientation was stated to underpin the study? e.g. grounded theory, discourse analysis, ethnography, phenomenology, content analysis | Positive-deviance framework | Methods |
| *Participant selection* | | | |
| 10. Sampling | How were participants selected? e.g. purposive, convenience, consecutive, snowball | Purposive sampling from a previously-completed survey | Methods |
| 11. Method of approach | How were participants approached? e.g. face-to-face, telephone, mail, email | Email | Methods |
| 12. Sample size | How many participants were in the study? | 37 | Results |
| 13. Non-participation | How many people refused to participate or dropped out? Reasons? | No one dropped out but we did not receive responses from everyone who was contacted with the recruitment email | -- |
| *Setting* | | | |
| 14. Setting of data collection | Where was the data collected? e.g. home, clinic, workplace | Interviewers were in the office (Ariadne Labs) and conducted interviews over the phone | Methods |
| 15. Presence of non-participants | Was anyone else present besides the participants and researchers? | One study team member was sometimes present to ensure proper audio recording of the interview | -- |
| 16. Description of sample | What are the important characteristics of the sample? e.g. demographic data, date | Table 1 provides characteristics of the facilities represented by the implementers who were interviewed<br>The number of states in which the facilities are located is provided in the text of the results. | Table 1<br>Results |
| *Data collection* | | | |
| 17. Interview guide | Were questions, prompts, guides provided by the authors? Was it pilot tested? | Yes; yes | Methods<br>Appendix |
| 18. Repeat interviews | Were repeat interviews carried | No | N/A |

| | | | |
|---|---|---|---|
| | out? If yes, how many? | | |
| 19. Audio/visual recording | Did the research use audio or visual recording to collect the data? | Yes, audiorecording | Methods |
| 20. Field notes | Were field notes made during and/or after the interview or focus group? | No | -- |
| 21. Duration | What was the duration of the interviews or focus group? | 60 minutes | Methods |
| 22. Data saturation | Was data saturation discussed? | Yes, saturation was achieved | Discussion |
| 23. Transcripts returned | Were transcripts returned to participants for comment and/or correction? | No | N/A |
| **Domain 3: analysis and findings** | | | |
| *Data analysis* | | | |
| 24. Number of data coders | How many data coders coded the data? | 3 | Methods |
| 25. Description of the coding tree | Did authors provide a description of the coding tree? | No | -- |
| 26. Derivation of themes | Were themes identified in advance or derived from the data? | Deductive coding was used | Methods |
| 27. Software | What software, if applicable, was used to manage the data? | NVivo | Methods |
| 28. Participant checking | Did participants provide feedback on the findings? | No | -- |
| *Reporting* | | | |
| 29. Quotations presented | Were participant quotations presented to illustrate the themes/findings? Was each quotation identified? e.g. participant number | Yes, quotes were provided to support the findings. Quotes are identified by the interviewees' type of facility (size of facility (small/medium, large) and level of implementation success (high, low) | Tables 2-4 |
| 30. Data and findings consistent | Was there consistency between the data presented and the findings? | Yes | Results and Discussion |
| 31. Clarity of major themes | Were major themes clearly presented in the findings? | Yes | Results Tables 2-4 |
| 32. Clarity of minor themes | Is there a description of diverse cases or discussion of minor themes? | Yes; the variation in implementation strategies included description of all variants (variants only mentioned by 1 respondent are indicated in Table 2). | Tables 2 |
